# Two Pools of ATP Detected in the Brains of Pediatric Patients with Myelin Oligodendrocyte Glycoprotein Antibody Disorders (MOGAD) by 3D ^31^P MR Spectroscopic Imaging (MRSI) at 7T

**DOI:** 10.1101/2022.01.13.22269241

**Authors:** Jimin Ren, Fang Yu, Benjamin M. Greenberg

## Abstract

Over the past four decades, ATP, the obligatory energy molecule for keeping all cells alive and functioning, was thought to contribute only one set of ^31^P MR signals in the human brain. Here we report for the first time the simultaneous detection of two pools of ATP in the human brain by high-resolution 3D ^31^P MRSI at ultrahigh field 7T. These two ATP pools differ in cytosolic Mg^2+^ concentration (1:0.5 ratio), with a resonance separation of 0.5 ppm at β-ATP, a well-established imaging marker of intracellular Mg^2+^ concentration. Mg^2+^ is a cofactor of ATPase and its deficiency is associated with immune dysfunction, free radical damage, perturbations in Ca^2+^ homeostasis, and development of atherosclerosis, dyslipidemia and a number of neurological disorders, such as cerebral vasospasm, stroke, migraine, Alzheimer’s disease, and Parkinson’s disease. Our study documents reduced Mg levels in the brain of patients with myelin oligodendrocyte glycoprotein antibody disorders (MOGAD), which is an idiopathic, inflammatory, demyelinating condition of the central nervous system (CNS) more common in pediatric patients. Low-Mg^2+^ ATP signals were detected mostly in the white matter regions in MOGAD, suggesting an association between Mg^2+^ deficiency and compromised functions of oligodendrocytes in maintenance and generation of the axonal myelin sheath. This preliminary study demonstrates the utility of the 7T 3D ^31^P MSRI for probing altered energy metabolism at reduced availability of Mg^2+^ rather than ATP itself. The potential correlation between [Mg^2+^] and disease progression over time should be assessed in larger cohorts.

**Author Approval:** Yes

## 1. Introduction

The brain is a heterogeneous organ consisting of distinct regions with varying cell compositions, metabolism and functions.^1,2^ ATP is the obligatory energy molecule for keeping all cells alive and functioning throughout the body.^3^ However, because of its intrinsically limited fuel reserves and exceedingly high energy demands, the brain is particularly susceptible to energy crises and development of various diseases.^4,5^ Brain energy homeostasis depends on the action of local neurometabolic and neurovascular couplings, which regulate various ATP-driven processes in response to neuronal activities, including increased blood flow to activated tissues to restore depleted ATP.^6,7^ Mg^2+^, the second most abundant intracellular cation, is the ATP activator in energy metabolism and also a co-factor in over 600 enzymatic reactions.^8^ Mg^2+^ deficiency has been associated with immune dysfunction, free radical damage, perturbations in Ca^2+^ homeostasis, and development of atherosclerosis, dyslipidemia, and a number of neurological disorders, such as cerebral vasospasm, epilepsy, stroke, migraine, anxiety, depression, Alzheimer’s and Parkinson’s diseases.^9,10^ Significantly decreased Mg^2+^ content has been found in autopsy samples of CNS white matter including demyelinated plaques in multiple sclerosis.^11^

^31^P MRS is a unique imaging modality capable of non-invasively measuring ATP and its energy metabolism in human brain.^12-17^ The triphosphate group of ATP molecule offers three characteristic, readily-observable ^31^P signals (α-, β-, and γ-ATP), which have found applications as endogenous imaging probes.^16-21^ Among these, β-ATP chemical shift can sense cellular Mg^2+^ with an upfield change indicating a reduction in Mg^2+^ level.^16,17^ On the other hand, γ-ATP can transfer its magnetization perturbation to inorganic phosphate (Pi) and phosphocreatine (PCr) by chemical exchange, a mechanism by which ATP fluxes can be estimated in the pathways of ATPase (Pi + MgADP → MgATP) and creatine kinase (PCr + MgADP ↔ MgATP + Cr).^18-21^

Since the first report of ^31^P MRS study in the human brain in the early 1980s,^22,23^ the brain is understood to yield only a single set of ATP signals. Observation of different pools of ATP, to the best of our knowledge, has not been directly evidenced in human brain. Resolving different ATP compartments may offer insights into fundamental clinical questions, such as why brain abnormalities (e.g., demyelination lesions) often attack certain brain regions and cellular components while sparing others, and what are the molecular basis that determines the onset of relapse, its time-course and variations between individuals.^24-29^

This study is carried out on a cohort of pediatric patients with myelin oligodendrocyte glycoprotein antibody disorders (MOGAD) − an idiopathic, inflammatory, demyelinating condition of the central nervous system (CNS), with heterogeneity in individual symptoms, radiological manifestations and clinical course.^30-32^ MOGAD lesions are often nonspecific on conventional MRI imaging; the diagnosis at the initial presentation requires an accurate serum antibody test.^31-33^ Currently, there is still a lack of understanding of the molecular, biochemical and cellular mechanisms that cause specific lesions to form, though several energy abnormalities are considered to be present at the sites of demyelination.^34-36^ Herein, we present evidence of the presence of two pools or compartments of ATP in this cohort, including one with remarkably reduced Mg^2+^ level. The cerebral heterogeneity in Mg^2+^ is revealed by high spatial resolution 3D ^31^P MRSI at ultra-high field 7T, with β-ATP as an endogenous imaging sensor.

## 2. Method

### 2.1 Protocol Approvals and Consent

This study was approved by the Institutional Review Board of The University of Texas Southwestern Medical Center. Seven participants with MOGAD disease between the age 11 and 21 years were recruited and informed and written consent obtained.

### 2.2 Data acquisition

The ^31^P MRSI data were acquired using a human 7T MRI scanner system (Achieva, Philips Healthcare), in combination with a ^31^P T/R birdcage volume coil of diameter 23 cm and length 10 cm (Gorten Center, Leiden University Medical Center, the Netherlands). The ^31^P coil was inserted into a cylindrical NOVA 1H T/R head coil for 1H shimming (second order PB-volume) and image planning. The patients were positioned head-first, and supine in the center of the ^31^P RF coil. The 3D ^31^P MRSI data were acquired at TR 0.5 s, in-plane resolution of 2×2 cm^2^ reconstructed to 0.9×0.9 cm^2^, slice thickness 2cm, k-space weighting (α = 1.7 and β = 1.0), and sampling points 4 k zero-filled to 8 k prior to Fourier transformation. Exponential apodization (20 Hz) was applied to FID data prior to Fourier transformation using the scanner software (SpectroView, Philips Healthcare). The chemical shifts of all ^31^P metabolites were referenced to PCr at 0 ppm. Cytosolic free Mg^2+^ was calculated based on chemical shift of the β-ATP, at the maximal peak magnitude or the weighted center of the peak, according to Lotti et al.^37^

To investigate whether the extra-cranial muscles of the head are a source of contamination to the observation of low-Mg ATP signal, comparative scans were conducted on both brain as well as calf muscles of five healthy subjects using a partial volume coil (Philips Healthcare) with a fully relaxed pulse-acquire sequence.

## 3. Results

### 3.1 β-ATP peak splitting: WM region

Fig.1A is a brain ^31^P spectrum summed over six consecutive voxels located in the left centrum semiovale. A total of nine major P-metabolites are visible, including PC, PE, Pi, GPE, GPC, PCr, ATP, NAD(H) and MM (a unspecified macromolecule). A unique feature of this spectrum is an identifiable β-ATP shoulder peak, appearing on the upfield side by a separation of 0.5 ppm (Fig.1A inset). Such a shoulder peak is not found at γ-ATP, whose chemical shift is less sensitive to Mg binding. However, the γ-ATP signal shows remarkable broadening, characterized by a linewidth more than two times broader than PCr. A similar peak broadening is also seen at Pi, whose chemical shift is known to be sensitive to pH, as compared to the more inert GPE and GPC signals nearby.

**Fig.1.**
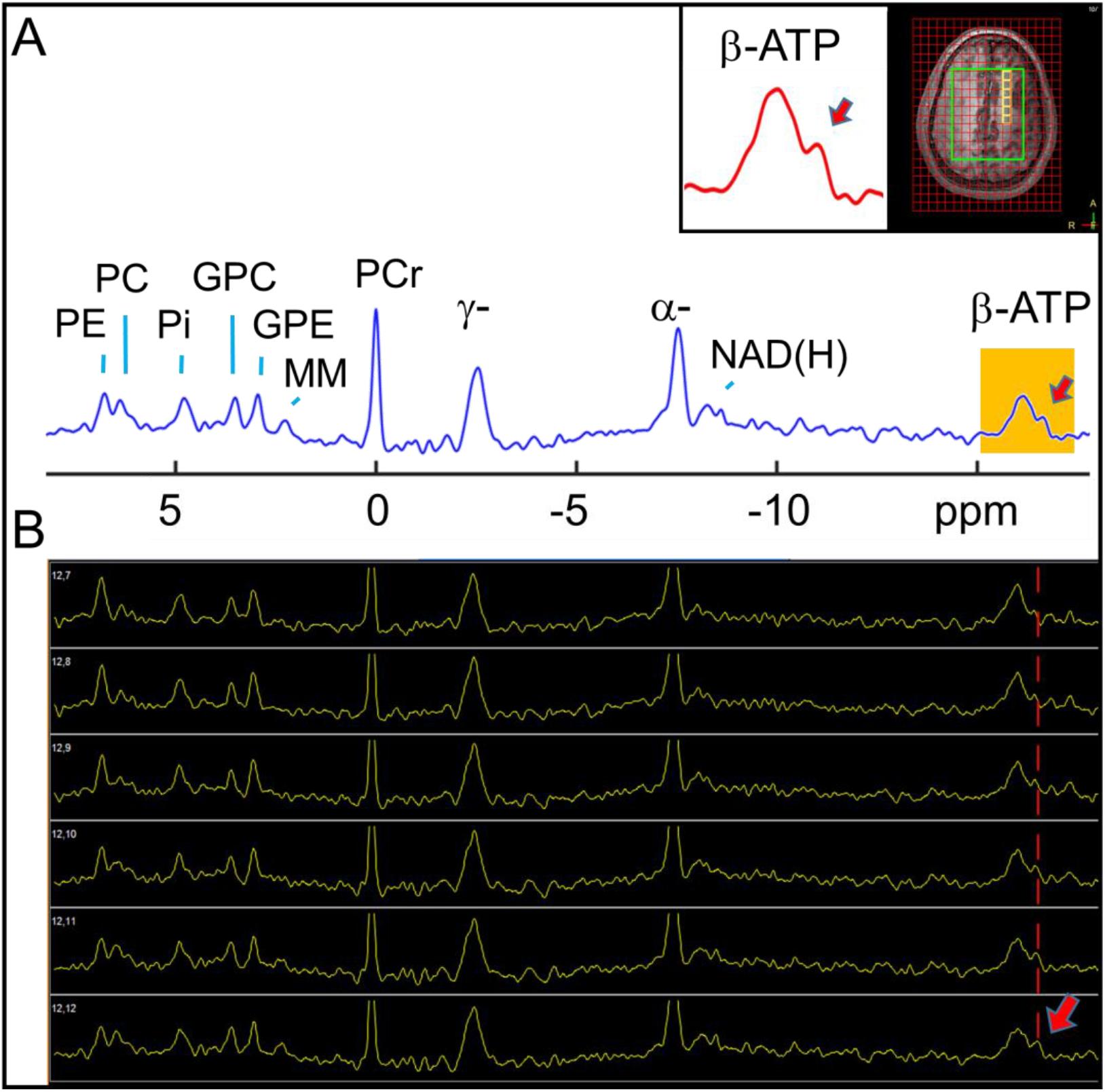
(A) A brain ^31^P spectrum summed over six consecutive voxels located in a subcortical WM-appearing region, showing the observation of two sets of β-ATP signals. (B) Scanner console screenshot image showing spectra from individual voxels (0.9×0.9×2 cm^3^). For better presentation of the low-magnitude β-ATP signal, the large PCr and α-ATP signals are truncated. Note the emerging of a separate β-ATP peak in the posterior voxels (bottom, arrow). Abbreviations: PE, phosphoethanolamine; PC, phosphocholine; Pi, inorganic phosphate; GPE, glycerolphosphoehtanolamine; GPC, glycerolphosphocholine; PCr, phosphocreatine; ATP, adenosine triphosphate; NAD(H), nicotinamide adenine diphosphate, a combination of the oxidized and reduced form; and MM, a unspecified macromolecule.

The splitting of β-ATP signal is observed not only in the voxel-summed spectrum (Fig.1A), but also in the spectrum of individual voxels (size 0.9×0.9×2 cm^3^). As shown in Fig.1B, the β-ATP shoulder peak becomes increasingly populated and hence more visible, as the voxel is located further posteriorly. The most posterior voxel of this dataset is characterized by a shoulder peak 25% of the size of total β-ATP.

Assuming that the two β-ATP peaks are from two separate compartments with different Mg^2+^ contents, then the cellular Mg^2+^ concentration is 0.18 ± 0.01 mM for the major pool and 0.09 ± 0.01 mM for minor upfield pool, estimated from the corresponding chemical shifts.

### 3.2 β-ATP peak splitting: deep brain region

Fig.2 shows the β-ATP resonance in a whole MRSI image plane. The data offers an overview of β-ATP splitting and its location-dependence. In general, as the voxel is located further inferiorly (caudally) in the brain, its β-ATP shoulder peak tends to be more populated, and more visually separated from the main one. The largest separation (0.5 ppm) is found in the most inferior voxels along the bottom row (Fig.2B, marked by arrows) where the two β-ATP components are approximately equal in size. The estimated cellular Mg^2+^ concentration is 0.20 mM and 0.10 mM, respectively, for these two ATP pools.

**Fig.2.**
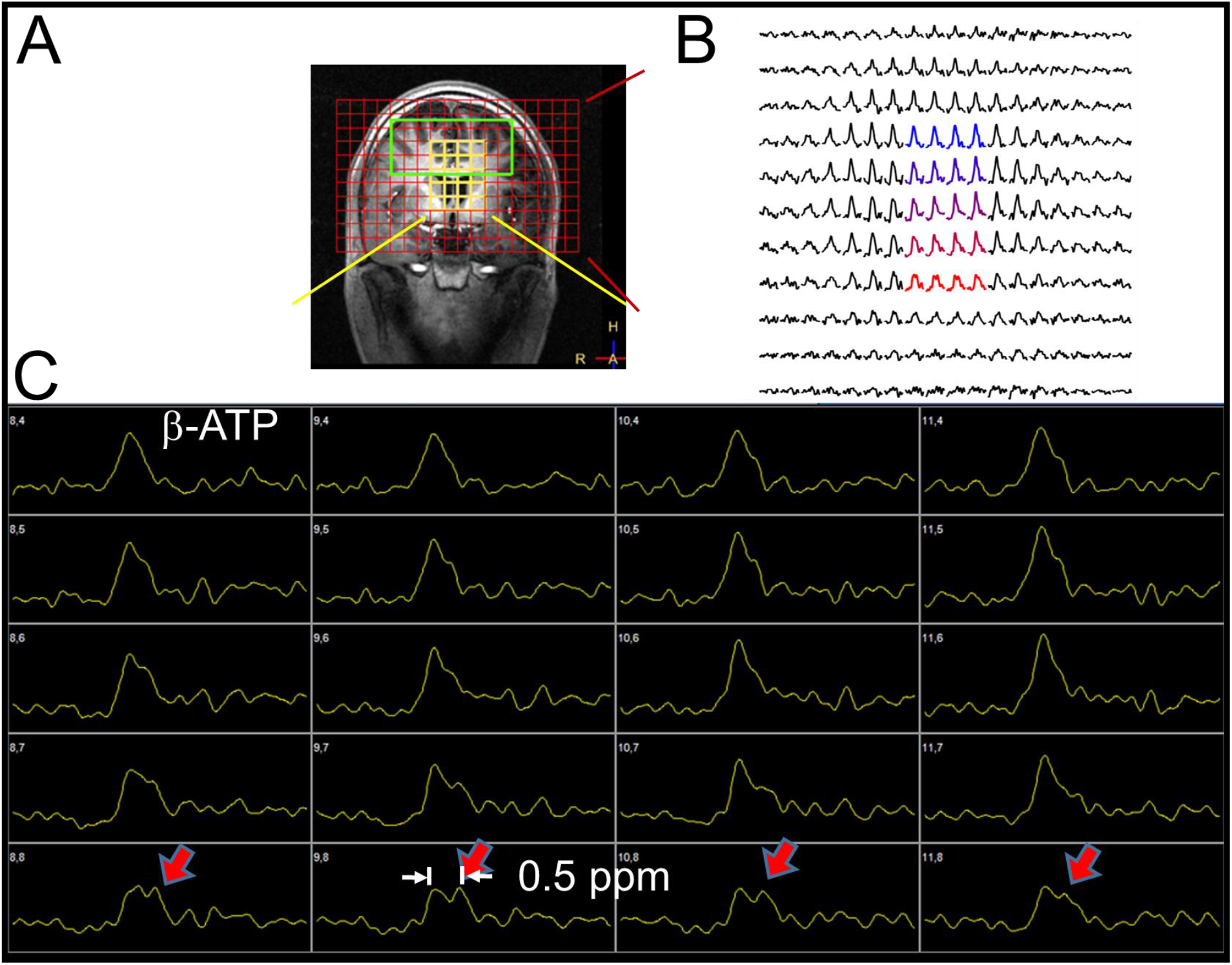
(A) A coronal MRI image with an overlay of the MRSI voxel matrix. (B) β-ATP signals from all voxels in the image plane. (C) Scanner console screenshot showing the enlarged β-ATP signals from the central 4×5 voxels. Note the clear separation of two β-ATP signals (marked by arrows).

### 3.3 Slant and flat-top β-ATP peaks

Fig.3A shows single-voxel spectra from consecutive voxels in the parietal region where β-ATP gives rise to a slant peak, instead of a symmetric peak or two separate peaks. Here both γ- and β-ATP resonances are featured with a peak slanted toward the upfield side, in contrast to PCr, which has a sharp and symmetric lineshape. This suggests that the slant peak is a result of two separate but not fully resolved ATP components, rather than an artifact. It appears that the ATP peaks are more slanted in the further inferior voxels with a greater portion of WM, compared to the superior voxel with a greater GM proportion. Under a homogenous model with a single ATP pool, the apparent intracellular Mg^2+^ concentration ranges from 0.18 mM in the superior voxel to 0.10 mM in the inferior voxel, estimated from the weight center of the β-ATP chemical shift.

**Fig.3.**
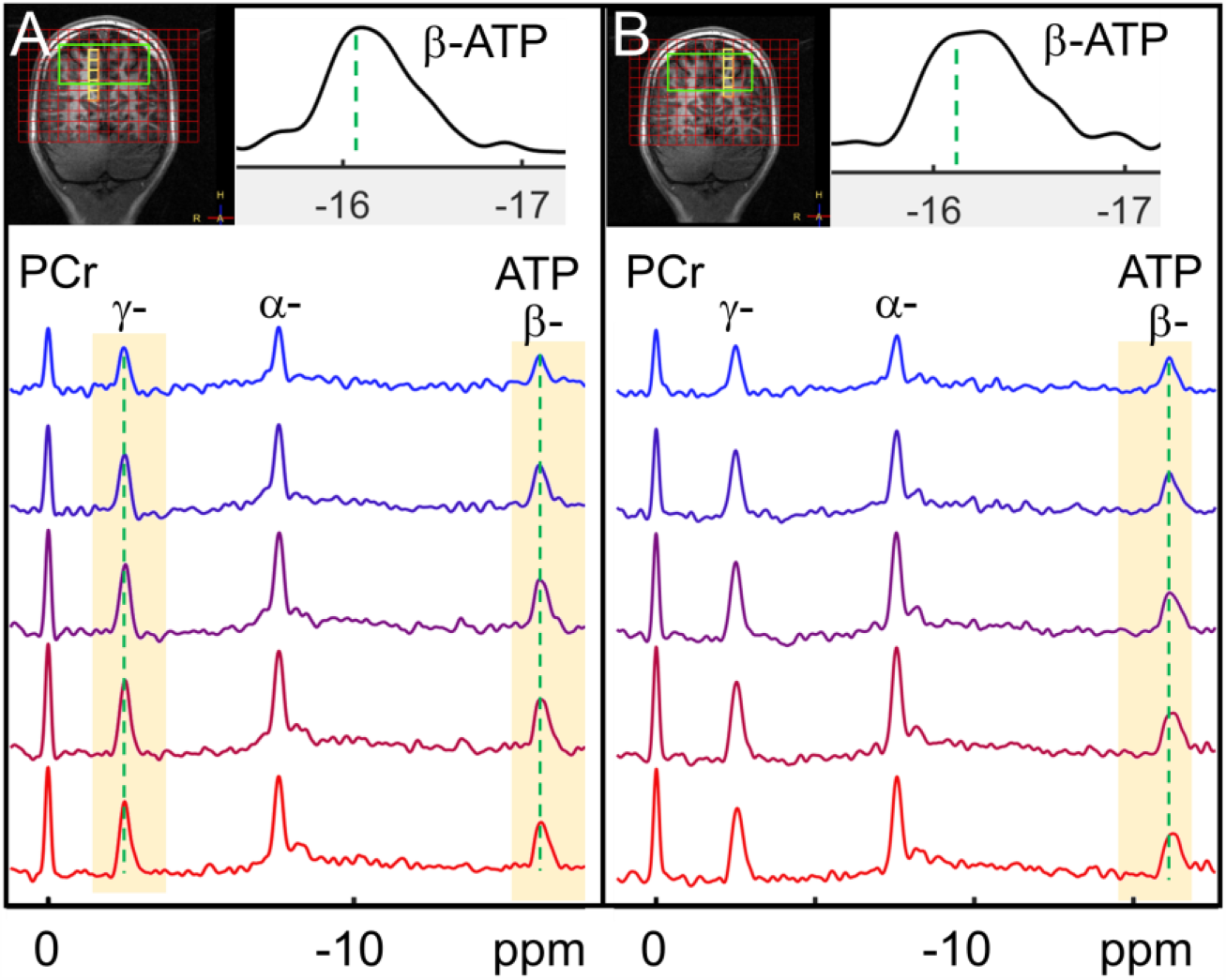
(A and B) ^31^P MRSI spectra from consecutive voxels selected in the anterior parietal lobes (A: right hemisphere; B: left hemisphere), to show the observation of slant and flat-top β-ATP peaks. The green dashed lines indicate the central position of the ATP peaks in the superior cortical voxels (top row, as reference). Note that the weighted center of the slant and flat-top β-ATP peaks is shifted toward upfield relative to the reference (insets).

Fig.3B shows the corresponding single-voxel spectra in the counter-hemisphere. Here flat-top β-ATP peaks, indicating comparable population between the low-Mg pool (0.09 mM) and the high-Mg pool as seen in the superior GM voxel (0.18 mM, top voxel), are observed in those inferior subcortical voxels with more WM involvement.

### 3.4 Contrast between cortical and subcortical tissues

Fig.4 shows two regional ^31^P spectra, obtained by summation of a large number of voxels selected from subcortical and outer cortical brain regions, respectively. These two regions have a very different distribution profile on PDE, as compared to PME and ATP (Fig.4A). It can be seen that, in both regions, all major peaks appear to be reasonably symmetric, except the subcortical β-ATP, which has a blunt shoulder peak visible upon enlargement (Fig.4B). Under the single pool model, the average intracellular [Mg^2+^] is estimated to be 0.14 mM in subcortical voxels, lower than that found in cortical voxels (0.20 mM). In parallel, a lower pH is found in the cortical tissue (pH6.95) than in the cortical tissue (6.99), measured by the Pi chemical shift in reference to PCr.

**Fig.4.**
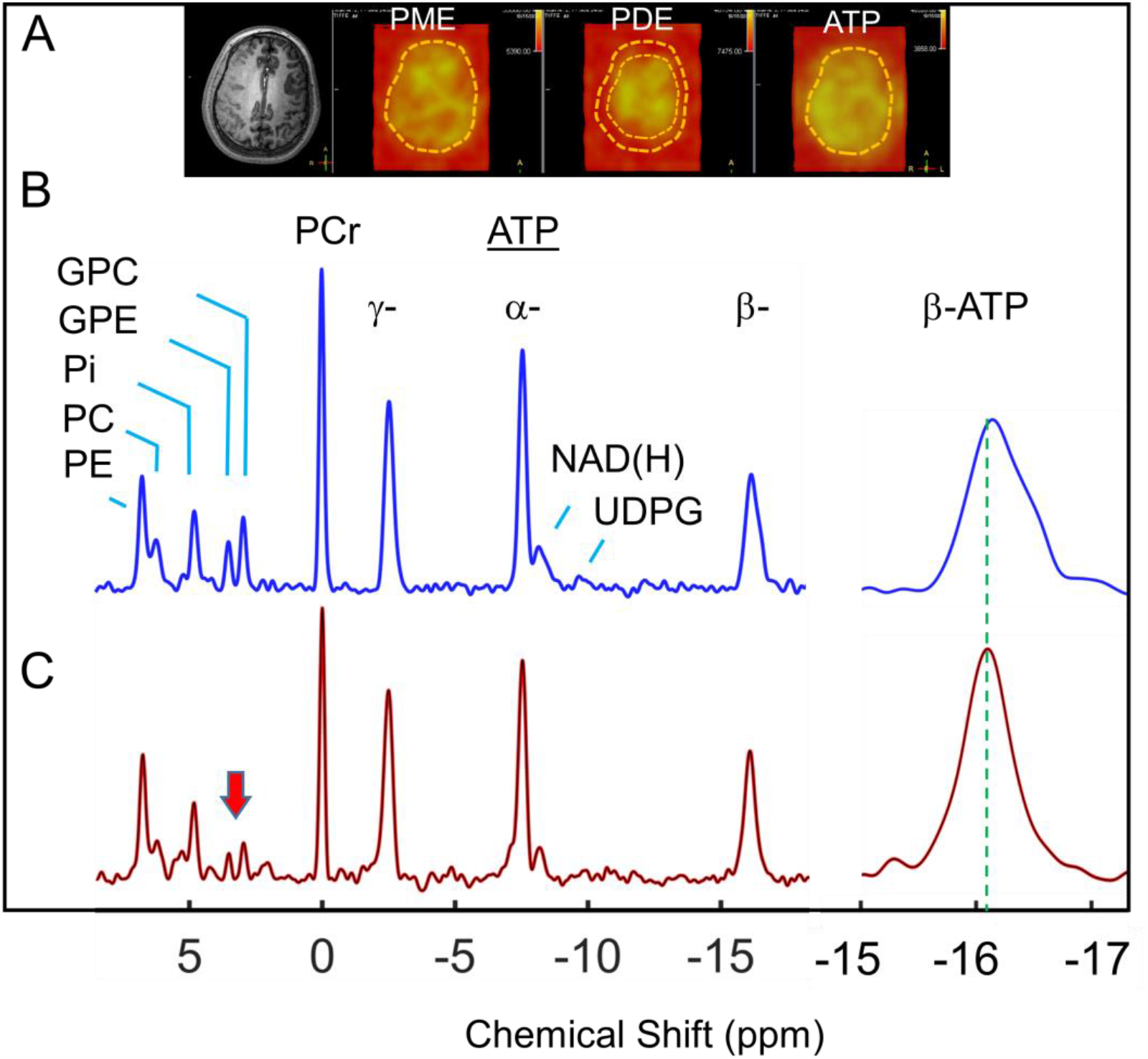
(A) ^31^P MRSI images for PME (PE + PC), PDE (GPE + GPC) and ATP. Note the PDE image contrast between outer cortical and inner subcortical regions. (B and C) ^31^P MR spectra summed over voxels selected from subcortical region (B: blue trace) and outer cortical region (C: red trace). The spectra are baseline-corrected and scaled at the magnitude of γ-ATP. The green dash line marks the central position of the cortical ATP peak. Note the reduced PDE signals in the cortical spectrum (red trace, arrow) and the broadened, upfield-shifted β-ATP shoulder signal in the subcortical spectrum (blue trace). Abbreviations: PME, phosphomonoester (= PE + PC); PDE phosphodiester (= GPE +GPC); and UDP(G), uridine diphosphate glucose and its drevatives.

### 3.5 Comparison between brain and muscle spectra

To investigate whether the β-ATP shoulder peak observed in the subcortical brain region is a contamination from the extracranial muscles of the head, ^31^P spectra were acquired from both the brain and skeletal muscle under the same conditions (RF coil and pulse-acquire sequence), as shown in Fig.5. The peak alignment reveals that, there is indeed a difference in β-ATP chemical shift between these two tissues. However, in reference to the chemical shift in the brain, the muscle β-ATP resonates on the downfield side, rather than upfield. This finding confirms that the β-ATP shoulder peak observed in the brain subcortical region is not a result of incidental contamination from the muscles of the head.

**Fig.5.**
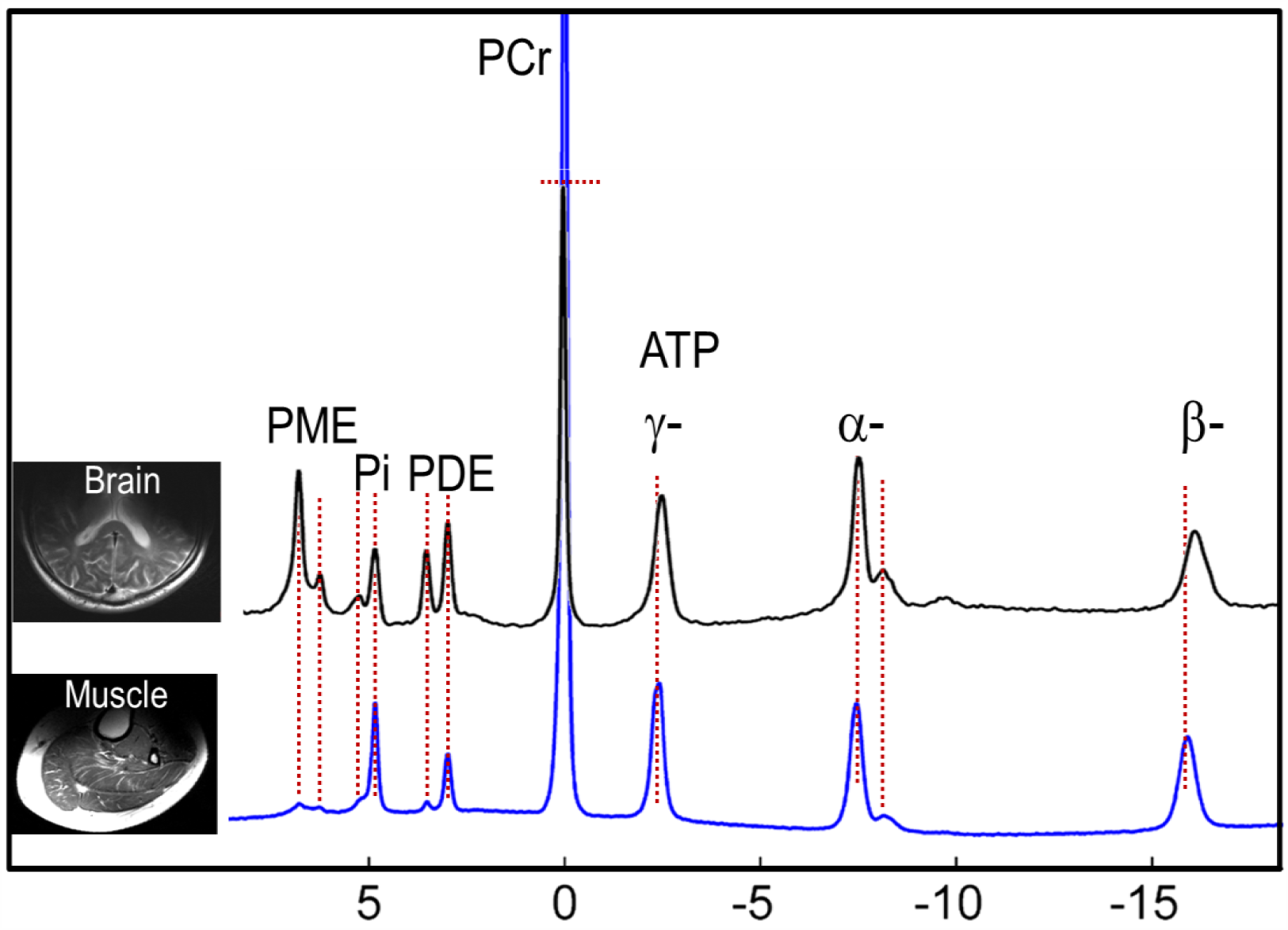
Cohort-averaged (n = 5) ^31^P MR spectra acquired from the brain (black trace) and the calf muscle (blue trace) under full relaxed condition with pulse-acquire sequence using the same partial-volume 1H/^31^P coil. Note the muscle β-ATP signal falls on the downfield side of the brain β-ATP signal.

## 4. DICUSSION

### 4.1 ATP splitting

In this study, two pools of ATP are detected directly in the brains of MOGAD patients by high-resolution 7T ^31^P MRSI (Figs.1-3). ATP peak splitting occurs mostly at β-ATP and with relatively high prevalence in the white matter. A maximal peak separation of 0.5 ppm is found in cases where the two pools have comparable ATP populations (Fig.2). However, there is a high spatial heterogeneity in peak separation, and an insufficient peak separation results in a slant, flat-top or shoulder β-ATP peak, with the weighted center of β-ATP markedly shifted toward upfield, indicating reduced local Mg levels. The spectral featured documented here may help identify similar patterns in future ^31^P MRS and MRSI studies, providing information regarding Mg^2+^ deficiency which may support the disease diagnosis.

### 4.2 Assignment considerations

#### 4.2.1 Extracellular ATP

It has been documented in the literature that ATP in the brain can exist in different compartments beside the intracellular space.^38,39^ Extracellular ATP has been detected in the interstitial space and the CSF by other more sensitive methods only feasible for interventional or in vitro studies. However, neither of these two compartments have sufficient ATP abundance to be observable by in vivo ^31^P MRS technique (with a detection limit of ∼ 0.02 mM).^40,41^ Separate ATP pools have not been resolved previously by non-localized and low-resolution MRS, likely due to a lack of sufficient spatial and spectral resolution, as most brain 31P MRS studies so far have been conducted at lower magnetic field strengths and/or on healthy subjects.

#### 4.2.2 Artifacts and muscle contamination

The appearance of the β-ATP shoulder peak is unlikely a result of random motion artifacts or spectral noise, since the shoulder peak has consistent resonance position across different brain regions and individuals, with a magnitude far above the spectral baseline noise (Figs. 1 and 2). The possibility that the shoulder β-ATP peak is an artifact from poor shimming and muscle contamination can also be excluded, given that the coexistent PCr signal, which is most sensitive to such artifacts, was found to be sharp, symmetric and base-clean (Figs.1-4). Even though PCr resonates at the same chemical shift between the brain and muscle, the muscle β-ATP contamination is unlikely to yield an upfield β-ATP shoulder peak in a brain spectrum (Fig.5).

#### 4.2.3 ^31^P-^31^P J-coupling

While ATP homonuclear ^31^P-^31^P J-coupling is detectable in the brain at lower fields (1.5T and 3T)^22,42^, the splitting is typically unresolvable at ultra-high fields, due to the line-broadening from the shortened T2.^43^ Indeed, none of the brain ATP resonances is featured with the typical pattern of J-coupling (α-: doublet; β-, triplet; and γ-, doublet^44,45^) in this 7T study, despite good shimming quality and high SNR (Figs.4 and 5). Furthermore, the inter-peak separation observed at β-ATP (0.5 ppm) is four-fold greater than that expected from a typical ^31^P-^31^P J-coupling constant under in vivo condition (15 -17 Hz^44,45^ or 0.12 – 0.14 ppm at 7T). Given such a large separation, as well as the use of a hard excitation pulse with FID-rather than echo-based data acquisition method, it seems unlikely that the β-ATP shoulder peak is a result of J-modulation effect.^44^

#### 4.2.4 MgATP vs other species

In healthy human brain, the majority of intracellular ATP is bound to Mg^2+^ in 1:1 MgATP complex,^17^ with Mg^2+^ chelated by two oxygen atoms of the β-phosphate group alone. This binding mode is favorable not only for rapid cleavage of the Pγ-O bond (removal of terminal γ-ATP group) in phosphotransferases and the Pα-O bond by nucleotidetransferases,^45^ but also for fast exchange of ATP between the free and the Mg^2+^-bound states without major alteration of ATP conformation (as compared to other chelating modes involving two different phosphate groups). The fast exchange leads to a Mg concentration-dependent change in ATP chemical shifts, with a much higher sensitivity at β-ATP than at γ- and α-ATP.^45^ The appearance of two β-ATP peaks is unlikely a result of slow exchange between the free and the Mg^2+^-bound ATP, as it would generate a much larger peak separation (2.3 ppm^44,45^) than that observed here (0.5 ppm). The possibility of ADP contribution can also be excluded since the β-ATP resonance, unlike γ- and α-ATP, which have very similar chemical shifts as β- and α- ADP, is free from ADP contamination.

It should be noted that, for easy communication, the triphosphate peaks are referred to as ATPs in this work, a strict assignment should be nucleoside triphosphates (NTP), of which ATP is the dominant species but unresolvable by 31P MRS from other NTP species, including guanosine-based GTP, uridine-based UTP and cytidine-based CTP. The urine and pyrimidine bases have no effect on the binding of Mg^2+^ to the phosphate groups.^45^

#### 4.2.6 pH effect vs Mg binding

In this study, assessed from Pi chemical shifts, a lower pH value is found in WM regions (pH 6.95) where a reduced magnesium is observed, as compared to the superficial GM regions (pH 6.99, Fig.4). The question arises as to whether the β-ATP shoulder peak is due to a lower intracellular pH rather than a lower Mg^2+^ concentration, as a reduction in pH, like Mg^2+^ binding to ATP, also induces an upfield shift at β-ATP. If this is the case, one would expect even a larger upfield shift at γ-ATP than β-ATP.^45-47^ Apparently, such a trend was not observed in our MOGAD cohort, indicating that a lower pH value in WM tissues is not a primary factor causing the appearance of the ATP shoulder peak.

### 4.3 Mg heterogeneity

The observation of location-dependent β-ATP resonance patterns, including symmetric, slant, flat-top, shoulder and separate β-ATP peaks, reflects spatial heterogeneity in Mg distribution in the brains of MOGAD subjects. Mg heterogeneity has been reported previously in skeletal muscle (a higher Mg content seen in gastrocnemius than soleus) by ^2^J_β-γ_(ATP) assessment,^48^ and recently in the brain of cancer patients by δ(β-ATP) measurement (a higher Mg observed in cancer lesions than distal tissues, from a single β-ATP peak).^49^ Of note, such direct evaluation of tissue Mg^2+^ and distribution cannot be replaced by blood samplings, given that a normal plasma Mg^2+^ may co-exist with a low tissue Mg^2+^ and that there is a lack of association between tissue Mg^2+^ and the Mg^2+^ level in erythrocytes which is under genetic control.^9^ Therefore, clinical studies need to take into account the tissue heterogeneity in Mg and other metabolic indexes such as pH, PCr, and PDE. ^49-51^

### 4.4 Mechanism, significance and implications

The observation of a maximal of β-ATP upfield shift (0.5 ppm) in MOGAD may lend support to the note that the CNS concentration of Mg^2+^ appears to have a critical level below which neurologic dysfunction occurs.^52^ The finding of low Mg^2+^ levels in white matter regions may suggest that oligodendrocytes, which are primarily responsible for the maintenance and generation of the axonal myelin sheath, are susceptible to Mg^2+^ deficiency. As cellular energetics is crucially dependent on the availability of the co-factor Mg^2+^, an adequate Mg^2+^ supply may play a protective role in WM damage. Given that several subunits of the Mg^2+^-dependent, glutamatergic NMRA receptors are expressed in oligodendrocytes,^52^ it can be speculated that Mg^2+^ could contribute to WM integrity by blocking NMDA receptors involved in excitotoxic damage. Mg^2+^ as a blocker of sodium and calcium entry can protect axons from nitric oxide-mediated degeneration.^53^ Mg^2+^ may also provide cellular protection via neuro-glial and vascular effects, including Mg^2+^-induced antioxidant actions, release of NO to induce vasodilation for improving blood flow and reducing vasospasm, and regeneratation of ATP for keeping neurons survive through ischemic insults.^54^ It should be emphasized, despite a normal total ATP level (i.e. without ATP depletion), Mg-deficiency (at reduced level of the activated energy form MgATP) may compromise the function of cellular ATP energetics. However, an Mg-dependent energy deficit may be reversible upon repletion of cellular Mg^2+^ in the affected tissues. This remains to be investigated with MOGAD.

## 5. Conclusion

We demonstrated that two sets of cerebral β-ATP signals can be detected by 3D ^31^P MRSI at 7T in MOGAD. One of the ATP pools with reduced cellular Mg^2+^ is found in WM tissues. The Mg-deficiency, if confirmed independently, may help develop clinical strategies to treat MOGAD, a newly recognized disease entity about which there is still a lack of imaging biomarkers and of understanding of the molecular mechanism of the pathogenesis. The ^31^P MRSI spectral features documented in this study may help future studies to identify sub-clinical effects of disease-modifying treatments at an earlier stage than is possible using measures based on clinical disease activity. Further investigation is needed to explore the origin of the Mg deficiency, the link between regional Mg levels and disease presentations, and longitudinal follow-up of disease progression with larger cohorts.

## Data Availability

All relevant data in the present work are contained in the manuscript.

## Declarations

I confirm all relevant ethical guidelines have been followed, and any necessary IRB and/or ethics committee approvals have been obtained.

## Data Availability Statement

All relevant data in the present work are contained in the manuscript.

## Clinical Protocols

IRB approved – NCT03942952 (PEDIATRIC SONICS: Pediatric Study of Neuropsychology and Imaging in CNS Demyelinating Syndromes)

